# Genetic surveillance reveals low, sustained malaria transmission with clonal replacement in Sao Tome and Principe

**DOI:** 10.1101/2024.07.15.24309968

**Authors:** Ying-An Chen, Peng-Yin Ng, Daniel Garcia, Aaron Elliot, Brian Palmer, Ronalg Mendes Costa d’ Assunção Carvalho, Lien-Fen Tseng, Cheng-Sheng Lee, Kun-Hsien Tsai, Bryan Greenhouse, Hsiao-Han Chang

## Abstract

Despite efforts to eliminate malaria in Sao Tome and Principe (STP), cases have recently increased. Understanding residual transmission structure is crucial for developing effective elimination strategies. This study collected surveillance data and generated amplicon sequencing data from 980 samples between 2010 and 2016 to examine the genetic structure of the parasite population. The mean multiplicity of infection (MOI) was 1.3, with 11% polyclonal infections, indicating low transmission intensity. Temporal trends of these genetic metrics did not align with incidence rates, suggesting that changes in genetic metrics may not straightforwardly reflect changes in transmission intensity, particularly in low transmission settings where genetic drift and importation have a substantial impact. While 88% of samples were genetically linked, continuous turnover in genetic clusters and changes in drug-resistance haplotypes were observed. Principal component analysis revealed some STP samples were genetically similar to those from Central and West Africa, indicating possible importation. These findings highlight the need to prioritize several interventions such as targeted interventions against transmission hotspots, reactive case detection, and strategies to reduce the introduction of new parasites into this island nation as it approaches elimination. This study also serves as a case study for implementing genetic surveillance in a low transmission setting.

## Introduction

Effective interventions have significantly reduced the malaria burden in the Democratic Republic of Sao Tome and Principe (STP), an island nation in the Gulf of Guinea, Central Africa ^1, 2, 3^. In recognition of the substantial progress achieved, the World Health Organization (WHO) has included the country in the E-2025 initiative, aiming to eliminate malaria by the year 2025 ^4^. Despite being in a pre-elimination phase, the annual incidence rate has consistently remained around 10 cases per 1,000 people over 7 years. Moreover, there was a 46% increase in case numbers reported in 2022 ^5^. These setbacks pose challenges to achieving the E-2025 goal and underscore the necessity of understanding the genetic structure of residual malaria transmission in STP in order to tailor effective elimination strategies.

The main island, Sao Tome, consists of six administrative districts. The majority of cases occur in the capital district, Agua Grande, where approximately 40% of the population resides ^1^. Malaria cases are reported year-round, with a slight decline often observed during the dry season from June to early September ^1, 6^. Located about 300 km off the coast of Gabon, STP’s increase in international tourism and active trade facilitate potential parasite connectivity between STP and other countries in West and Central Africa ^7^. However, there has been limited exploration of the role of importation on malaria transmission in STP ^6^. Malaria interventions in STP have focused on vector control ^1, 8^, passive case surveillance ^6^, and/or mass drug administration ^9^, but have not included routine collection of travel history or performing reactive case detection, which may be useful strategies for preventing imported malaria and additional onward transmission in low-transmission settings ^10^.

The current understanding of malaria transmission in STP includes epidemiological findings, the evaluation of intervention effectiveness, and the analysis of specific genes in the local vector (*Anopheles coluzzii*) and the malaria parasite (*Plasmodium falciparum*) ^1, 6, 11, 12, 13^. However, a gap remains due to the lack of integration between epidemiological metadata and genomic information, which is crucial for understanding the structure of residual transmission in STP. With advancements and cost reductions in sequencing technology, an increasing number of countries are adopting genomic surveillance approaches to track transmission dynamics, potential importations, and the spread of drug-resistant parasites ^14, 15, 16, 17, 18, 19, 20, 21, 22, 23, 24, 25, 26, 27^. Several genetic metrics have been proposed to estimate malaria transmission, including the multiplicity of infection (MOI—which denotes the number of genetically distinct parasite strains within an individual), within-host relatedness, *F_ws_*, and pairwise relatedness between infections ^22, 28, 29, 30, 31, 32, 33^. Moreover, in settings with low transmission, genetic data can capture changes in circulating parasites that may not be easily detected through traditional epidemiological or clinical measures, thereby providing useful information for elimination efforts ^14, 34^.

To understand the genetic structure of residual malaria transmission in STP, we performed amplicon sequencing to obtain the first genome-wide genetic dataset of malaria parasites in the country. By integrating parasite genomic data with epidemiological metadata, this study offers spatiotemporal inferences on transmission clusters that can help guide effective malaria control strategies. The fine-scale analysis of genetic metrics and transmission inferences can serve as a valuable case study for applying genetic epidemiology to malaria in a low-transmission setting, particularly as more countries experience transmission declines and approach malaria elimination.

## Materials and Methods

### Ethics statement

The transfer, shipment, and use of malaria dried blood spots (DBS) and encrypted case surveillance data for research analysis in Taiwan was approved by the Centro Nacional de Endemias (CNE) in STP (reference no. OF°N°20/P°CNE/2016). This study was reviewed and approved by the Research Ethics Committee of National Taiwan University Hospital (reference no. 201110023RD) and the Research Ethics Review Committee of National Tsing Hua University (reference no. 11012HM135).

### Study materials

A total of 7,482 malaria dried blood spots (DBS) from the Central Hospital Ayres de Menezes (HAM) were obtained from a surveillance project conducted by the Taiwan Anti-Malarial Advisory Team in partnership with the Centro Nacional de Endemias (CNE) of STP (Fig. S1a). We randomly selected 1,629 DBS, evenly distributed across our study period nationwide, ensuring around 20 samples per month from 2010 to 2016. The average sample sizes by year in areas with relatively high and low malaria endemicity were 70 and 5 per district, respectively. The DBS, along with the basic characteristics of patients (including age, gender, onset date, residential district and village, treatment regime, and diagnostic results), were collected from patients who had either visited or reported to HAM and were confirmed positive for *Plasmodium falciparum* infections through microscopy and/or rapid diagnostic tests. The total numbers of samples from six administrative districts, Agua Grande (AG), Me-Zochi (MZ), Lobata (LO), Cantagalo (CT), Lemba (LE), and Principe (PR) were 936 (57%), 375 (23%), 236 (15%), 74 (5%), 6 (0.4%), and 2 (0.1%), respectively. Agua Grande is the capital district. The incidence data used in this study were provided by the Taiwan Anti-Malarial Advisory Team in STP ^1^. The data supporting the findings of this study have been deposited in the GitHub repository, accessible at https://github.com/hhc-lab/malaria_genetics_STP.

### DNA extraction and qPCR

DNA was extracted from DBS using the Tween-Chelex method described in Teyssier et al., 2021 ^35^. The 6-mm disc from the DBS punch underwent two-step washes with 0.5% Tween 20 in 1X PBS and 1X PBS at 4 °C. After removing the wash solution, the DBS disc was covered with 10% Chelex 100 resin in water at 95 °C for 10 minutes and centrifuged at 15,000 rpm for 10 minutes. The resulting supernatant, which contained the extracted DNA, was then transferred into a PCR tube and stored at −20 °C.

Parasite density was measured using the *varATS* qPCR protocol as previously described ^36^. DNA extracted from mock DBS with known parasite densities of 1, 10, 100, 1,000, and 10,000 parasites/μL served as qPCR standards. Duplicate sets of these standards were used to generate a standard curve for each run, with a linearity (*R*^2^) value exceeding 0.98 being considered a validated run. The parasite densities of the samples were then benchmarked using the established standard curve. A total of 1,478 samples were confirmed as positive with quantified parasite densities and were processed for amplicon sequencing.

### Amplicon sequencing

The amplification of two pools, comprising 165 diversity amplicons, 38 drug-resistance amplicons (containing 68 important drug-resistance sites), and 38 immune, diagnostic, and species-related amplicons, was obtained using the MAD4HatTeR V.3 protocol, building on a previously published method targeting diverse microhaplotypes ^37, 38^ and the CleanPlex library preparation kit from Paragon Genomics, CA, USA ^39^. The two pools were combined and processed through clean-up steps with magnetic beads for each sample. Samples were then pooled together and gel-extracted based on parasite density and the presence of primer dimers. All sample pools underwent quality checks on a TapeStation (Agilent, CA, USA) and were sequenced with 150 bp paired-end reads on Illumina sequencers.

For quality assurance, each sample plate included at least one positive control (3D7) and several negative controls randomly placed in each plate to monitor amplification efficiency and detect potential contamination. If the positive controls did not perform well (less than 100 reads per amplicon), or if the negative controls showed more than 10 reads per targeted amplicon on average, the entire plate was discarded and re-prepared until good-quality and contamination-free results were obtained.

### Bioinformatic pipeline and quality filtering

The amplicon data were processed through the allele calling pipeline V0.1.8 (available at https://github.com/EPPIcenter/mad4hatter). The pipeline consists of core modules designed to filter and correct demultiplexed reads, remove adapters, and enable accurate and precise identification of variant alleles. In this study, we focused on genetic diversity analysis using 165 diversity amplicons to estimate parasite genetic metrics, and seven drug-resistance amplicons to infer drug-resistance haplotypes.

To ensure data quality, we applied the following criteria to select high-quality samples for analyses using diversity amplicons: (a) total reads per sample exceeding the number of amplified amplicons multiplied by 100, (b) amplicon coverage exceeding 75%, and (c) more than half of the amplicons having 100 or more reads (Fig. S2). These standards ensured uniform and adequate amplification across most amplicons per sample. For drug-resistance amplicons, the criterion was that more than half of them should have 10 or more reads. A total of 980 samples met these requirements and were included in the analysis (Fig. S1b).

### Analysis of parasite genetic diversity

We utilized the *MOIRE V3.1.0* R package to implement a Bayesian approach for estimating MOI from polyallelic data, accounting for experimental error ^28^. We also estimated a new metric of diversity, the effective MOI (eMOI), by adjusting the true MOI for underlying within-host relatedness (*r*_*w*_), the average proportion of the genome that is identical by descent across all strains within the same host ^28^. The eMOI was defined as (MOI – 1) * (1 –*r*_*w*_) + 1. We used the means from the posterior distributions of MOI, eMOI, and *r*_*w*_ as their point estimates. A polyclonal infection was defined as having eMOI greater than 1.1.

We employed the *Dcifer V1.2.0* R package, an identity-by-descent (IBD) method, to estimate between-host relatedness from polyallelic data (*r*_*b*_) ^31^. Using the *MOIRE*-estimated MOI and allele frequencies as inputs, we obtained an estimation of between-host relatedness for each pair, ranging from 0 to 1, accompanied by a corresponding p-value for statistical significance. Genomic clusters were identified by applying various levels of 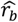, with 0.9 being the default and most stringent threshold and 0.3 being the least. The resulting 62 clusters using the 0.9 threshold were then ordered and named by their size (from small to large; C1 to C62). The clustering network was visualized, displaying only the links with a significance level that had a Bonferroni-corrected p-value of less than 0.05.

Given the seasonality of malaria transmission in STP, we classified annual seasons based on the cycle of transmission intensity (Table S1 and Fig. S3) instead of using the calendar year. The year was categorized from the beginning of the rainy season to the conclusion of the dry season, that is, from September of the prior year to August of the current year. Malaria incidence rates were also calculated based on the adjusted year and geographical groups (Capital or Others). Regression analysis was employed to identify the relationship between MOI, eMOI, and polyclonal infections and other characteristics, including the sampling year, season (rainy or dry), residential area (Capital or Others), location (Q1 and Q2, where Q1 represents locations at the village administrative level with malaria case numbers ranking in the top 25% within each district and Q2 represents others), age, gender, parasite density from qPCR (log_10_ transformed), and treatment regime (artemisinin-based combination therapy or quinine).

### Estimation of effective population size

We estimated the effective population sizes based on temporal changes in allele frequencies between consecutive years using the temporal method in *NeEstimator V2.1* ^40, 41, 42^. We used SNPs from diversity amplicons and excluded those with more than 50% missing data. Since the generation time for malaria parasites in STP was not previously estimated, we explored a range of values from 2 to 9 generations per year.

### Analysis of drug-resistance mutations

A total of 9 amino acid sites across 3 resistance genes (*mdr1*, *dhfr*, and *dhps*) were analyzed. For each sample, we first identified the predominant amino acid at each site using read counts, then combined the major amino acids from six sites in *dhfr* and *dhps* and three sites in *mdr1* (Table S2) to determine the drug-resistance haplotype for SP (sulfadoxine-pyrimethamine) and ASAQ/AL (artesunate-amodiaquine/artemether-lumefantrine), respectively.

### Principal component analysis (PCA)

We performed PCA for sequences from STP and six African countries, including Gabon, Nigeria, Ghana, Cameroon, Tanzania, and Kenya. Sequences from six African countries were obtained from the *Plasmodium falciparum* genomic variation dataset version 7 (Pf7) ^43^. For each sample, sequences in the regions corresponding to the genomic locations of the diversity amplicons from STP samples were extracted using *BCFtools* ^44^. These sequences were then merged with STP amplicon sequences to create a unified dataset. For STP samples with more than one sequence per amplicon per sample, the sequence with the highest read count was used. The concatenated sequences were aligned using the long sequence aligner *FAME* ^45^. Sites with homology greater than 90% and sites containing more than 50% gaps were removed. To perform PCA ^46^, aligned sequences were converted into a Pandas DataFrame, and bases were one-hot encoded using a base map.

## Results

### Genetic metrics were consistent with low malaria transmission but did not temporally track with incidence rate

With 980 sequenced samples collected nationwide over 7 consecutive years (Fig. S1b and Table S1), we examined four genetic metrics related to transmission intensity: MOI (multiplicity of infection), eMOI (effective MOI), within-host relatedness 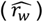, and the proportion of polyclonal infections. Overall, the MOI and eMOI among the samples were both low (mean = 1.3 [MOI] and 1.08 [eMOI]; Fig. 1a). Monoclonal infections were predominant, with polyclonal infections accounting for only 11% of samples (Fig. 1b). Among polyclonal infections, the average within host relatedness 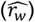 was high at 0.7 (Q1-Q3 = 0.6-0.8; Fig. 1c). Moreover, the estimated effective population sizes based on allele frequency changes are below 100 across all years (Fig. S4). Collectively, these data indicating low within-host diversity are consistent with low transmission intensity in STP during the study period. While these genetic metrics varied over time, their temporal trend differed from that of the incidence rate (Spearman’s rho < 0.6 and *p* > 0.2 for all; Fig. 1a, 1b). During the study period, the incidence was highest in 2012, while MOI was higher before and after that (2011 and 2014).

**Fig. 1.**
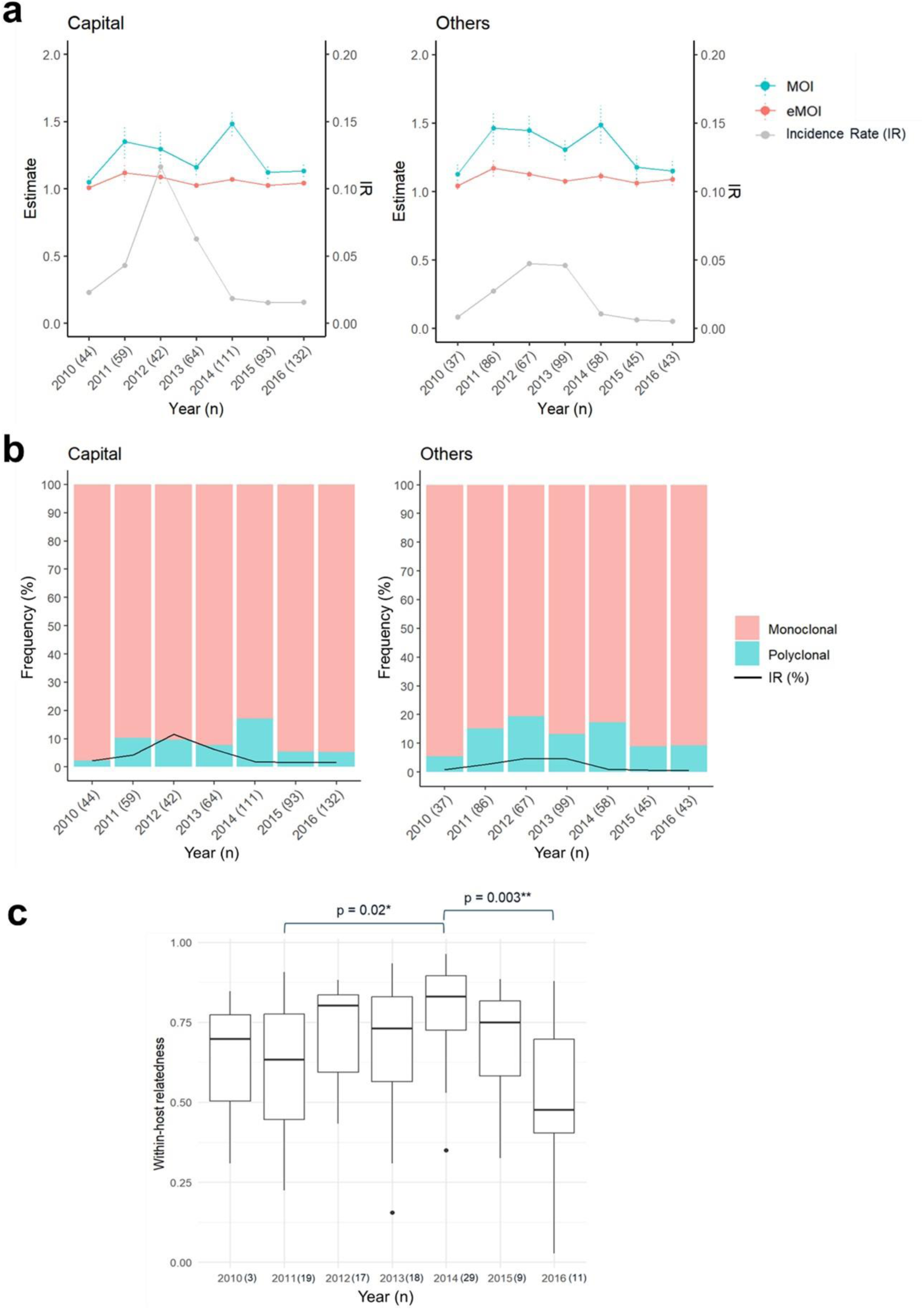
Temporal variation of parasite genetic metrics and malaria incidence rates. (a) MOI, eMOI, and incidence rates by reclassified year in the capital district (left) and other districts (right). The dots represent the mean for each year, and the bars for MOI and eMOI indicate the standard errors. The sample size for each year is shown in brackets on the x-axis. (b) Proportion of polyclonal (cyan) and monoclonal (pink) infections by year. Polyclonal infection is defined as eMOI greater than 1.1. (c) Boxplot of within-host relatedness (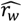) for polyclonal infections each year. The difference in within-host relatedness between years was tested using Dunn’s test.

In addition to temporal differences, we identified two other significant factors associated with the proportion of polyclonal infections, MOI, and eMOI (Table S3). A higher proportion of polyclonal infections (14% vs. 9%), along with higher MOI (1.34 vs. 1.24) and eMOI (1.10 vs. 1.05), were observed in locations outside or bordering the capital district compared to within the capital (Fig. 2 and Table S3). This suggests that effective recombination rates were higher outside or bordering the capital district, despite the fact that the capital district harbored the highest number of malaria cases. Another significant factor was age — individuals aged above 5 years exhibited a higher proportion of polyclonal infections compared to children under 5 (Table S3). This trend could be attributed to increased mobility among older individuals, leading to greater exposure to infectious sources. Additionally, a higher proportion of mild or asymptomatic infections among older individuals ^6, 47^ may result in delayed treatment, facilitating longer duration infections and the potential for superinfection.

**Fig. 2.**
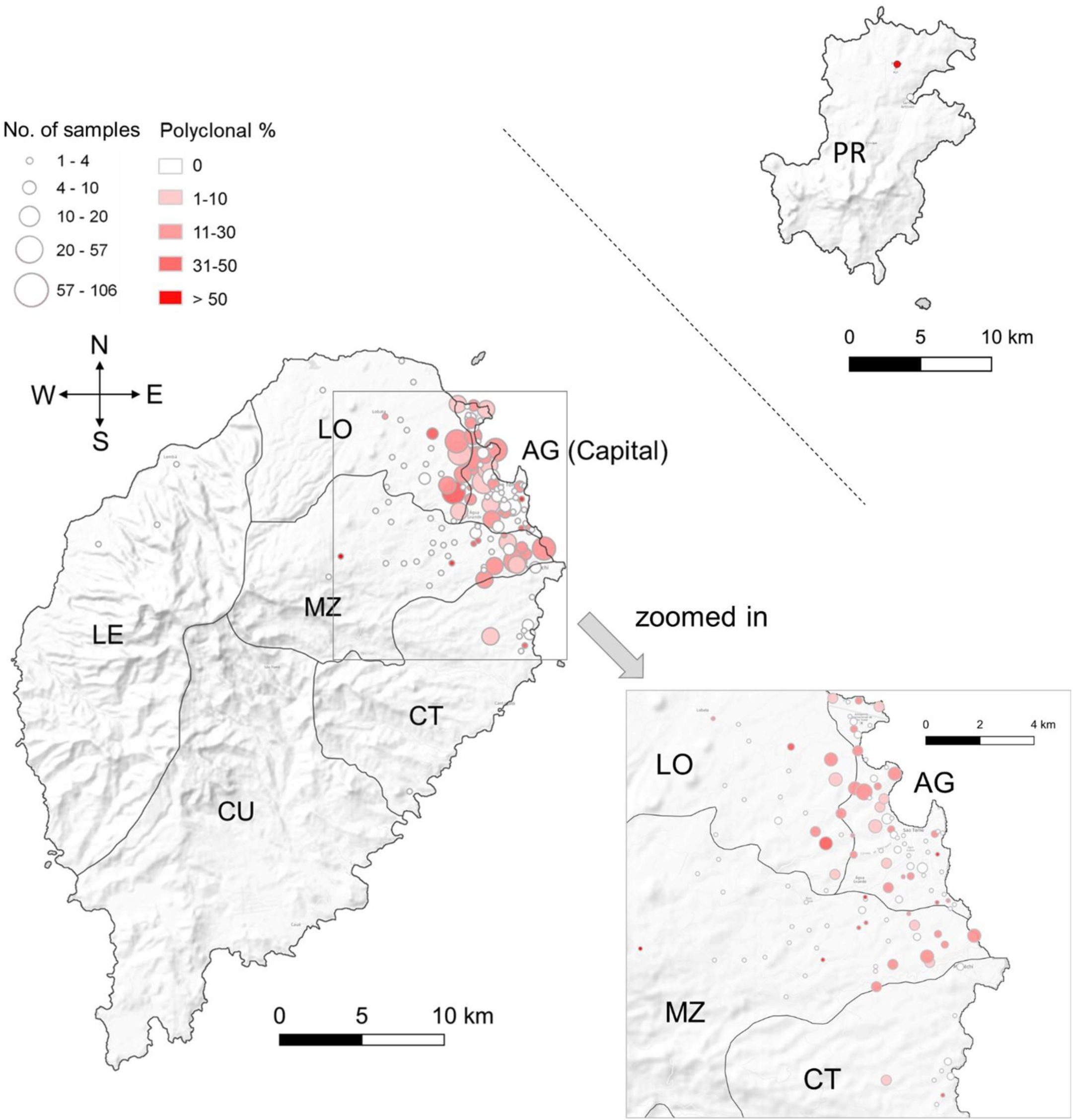
Distribution of sequenced samples and proportion of polyclonal infections. Sao Tome Island comprises six administrative districts: Agua Grande (AG, capital), Lobata (LO), Me-Zochi (MZ), Cantagalo (CT), Caue (CU), and Lemba (LE). Principe (PR) island is located 173 km away from Sao Tome Island. The dots on the map were plotted at the centroids of each location, which may represent residential hamlets, specific places, or roads reported by infected individuals. The size of each dot is proportional to the number of samples sequenced from that location, while the color gradients indicate the level of polyclonal infections.

### Genetic relatedness revealed sustained local transmission with continuous clonal replacement

A large majority of samples (88%) were infected by parasites closely related to at least one other sample (between-host relatedness 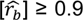), forming 62 highly related clusters (Fig. 3a). The high proportion of clustered samples and relatively small number of clusters identified each year (ranging from 8 to 31) suggest that our study captured the majority of a limited extent of population genetic diversity, despite only genotyping a representative sample of cases. In addition, these data suggest that most detected cases resulted from local transmission and, corroborating the incidence and within-host diversity data, are consistent with low transmission intensity. Despite low and seasonal transmission, the majority of clusters (63%) were found across multiple years, indicating sustained local transmission.

**Fig. 3.**
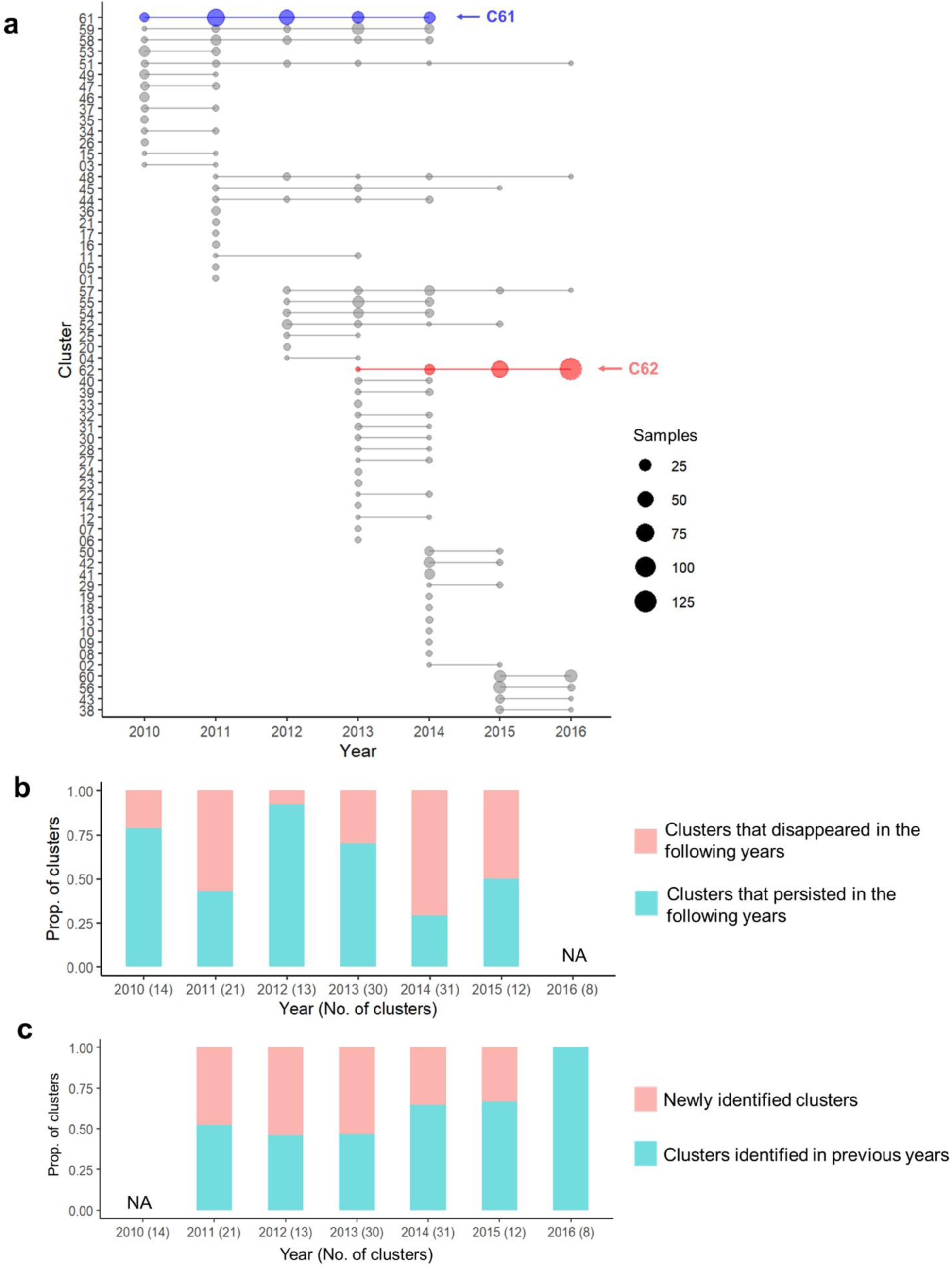
Characterization of clusters across years. (a) Using a cutoff of 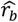 ≥ 0.9, 62 clusters were identified. The two most prevalent clusters are marked in blue (C61) and red (C62), respectively. (b) Proportion of clusters that disappeared in the following years (pink) and those that persisted (cyan). (c) Proportion of clusters that were newly identified in each year (pink) and those that were identified in previous years (cyan). NA = Not Applicable (since 2010 is the first year and 2016 is the last year, there is no information for previous years and following years, respectively).

While clusters frequently persisted over multiple years, few spanned the entire study period. Generally, we observed turnover in the parasite population over time (Fig. 3a). We found transitional changes in the parasite population over the years across a range of relatedness cutoffs from relaxed 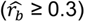 to stringent 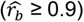 (Fig. 4a). Parasites collected in the same year exhibited higher relatedness compared to those collected further apart in time (Fig. 4ab).

**Fig. 4.**
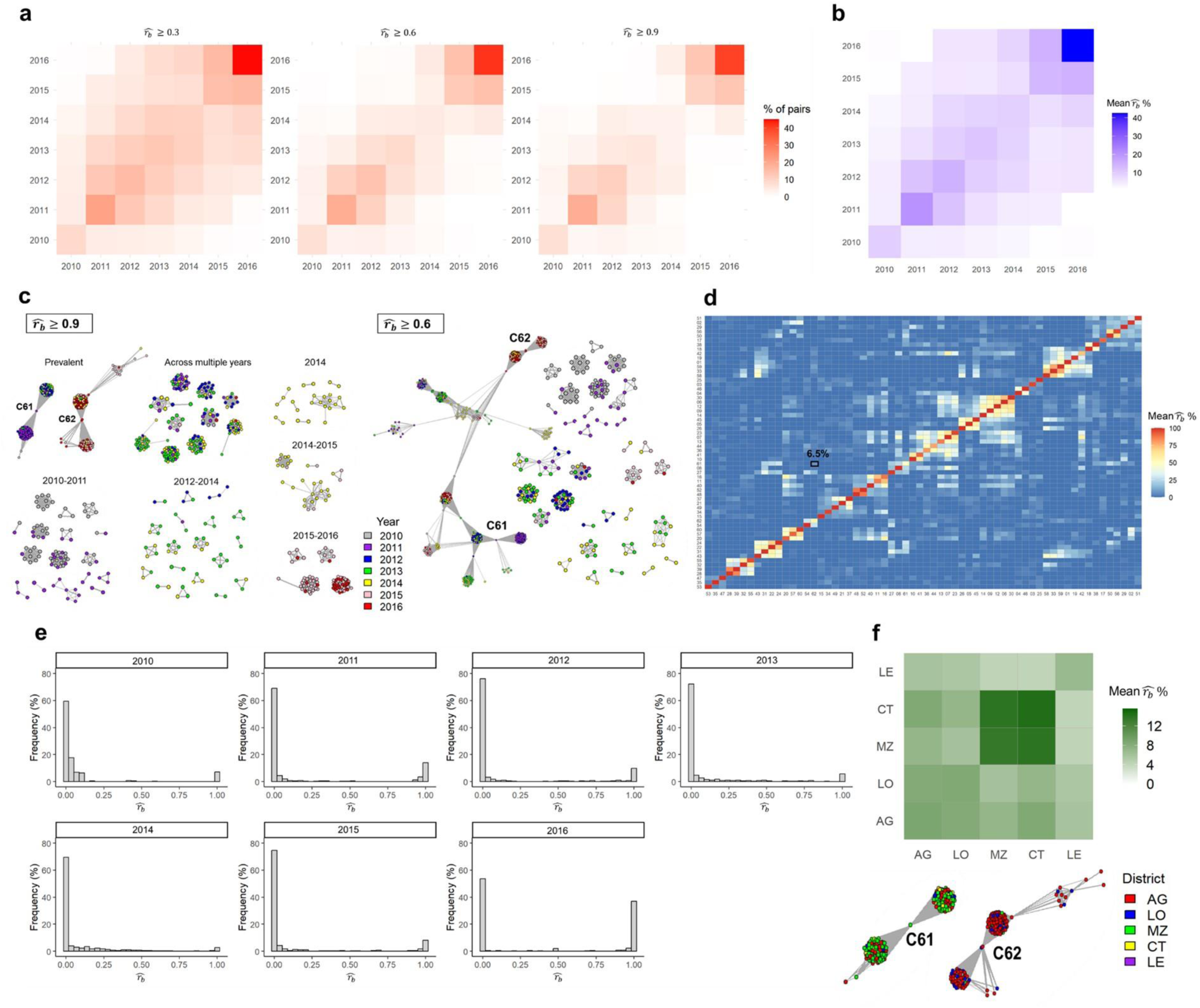
Estimated pairwise relatedness (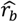) across time, space, and clusters. (a) Proportion of pairs with 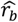 over 0.3, 0.6, or 0.9 between all pairs of years or within the same years. (b) Mean 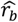 across and within years. (c) Samples with significant relatedness above 0.9 (left) and 0.6 (right) with a Bonferroni-corrected p-value of less than 0.05 are connected. Two predominant clusters, C61 and C62, are connected by 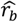 ≥ 0.6 through some transitional infections in 2013 and 2014. (d) Mean 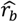 among 62 clusters identified using 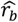 ≥ 0.9 cutoff. The average relatedness between C61 and C62 is 6.5% (marked in square). (e) Distribution of 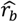 among samples within the same year. (f) Mean 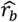 between districts in Sao Tome main island and the composition of districts in two predominant clusters, C61 and C62.

The mean within-year relatedness peaked in 2016 (average 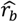 = 0.43), while the mean between-year relatedness between 2016 and earlier years, such as 2010 or 2011, was very low (average 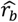 ≤ 0.02) (Fig. 4b). Drug resistance haplotypes also exhibited a temporal trend, with mutations suggesting a decreasing resistance level to SP (Fig. 5a) and an increasing resistance level to AL (Fig. 5b).

**Fig. 5.**
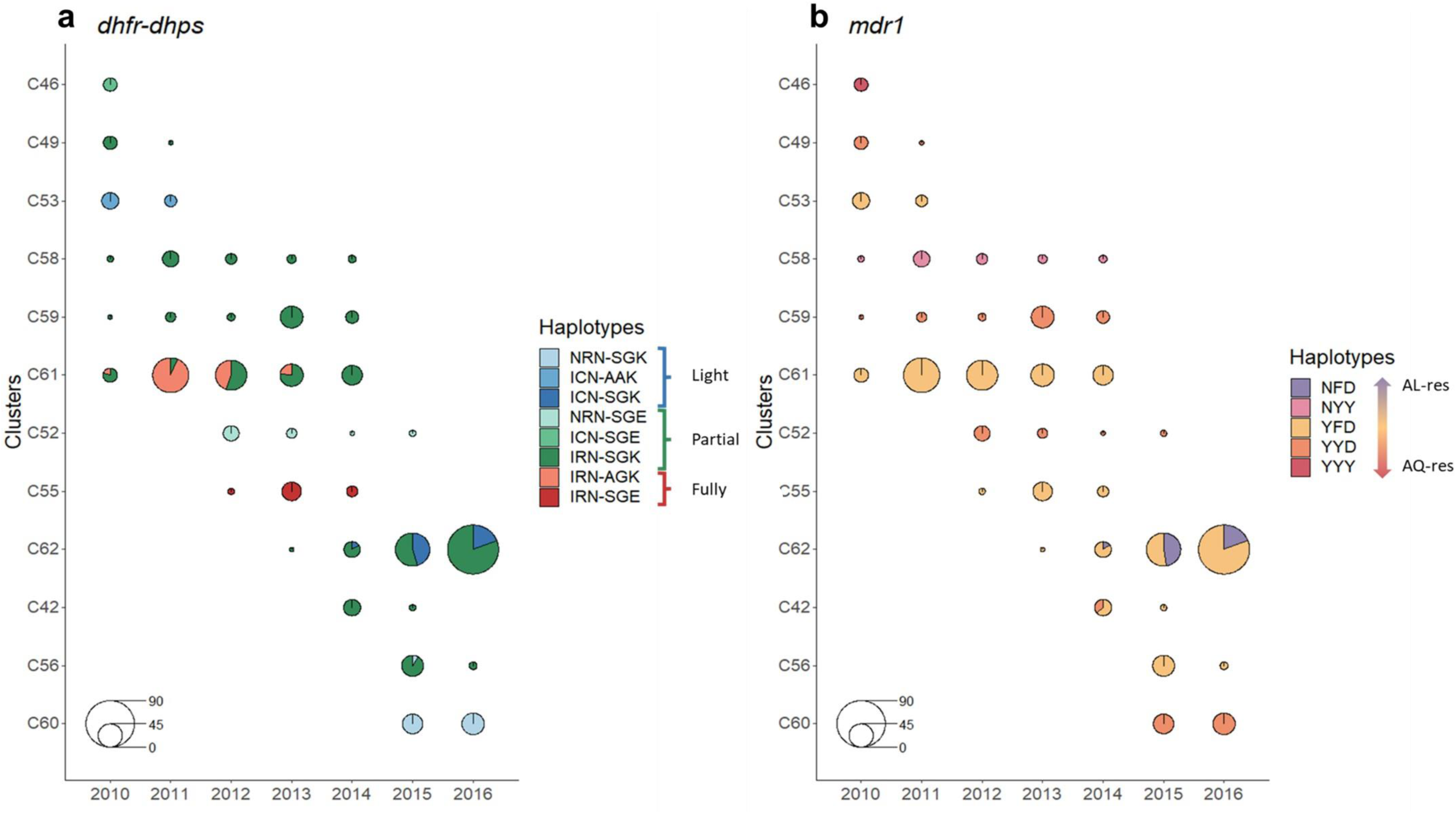
Haplotype changes in (a) *dhfr*-*dhps* and (b) *mdr1* over the years. The *dhfr*-*dhps* haplotypes (N51I/C59R/S108N-S436A/A437G/K540E) are color-coded according to their resistance levels: blue for light resistance (3 mutants), green for partial resistance (4 mutants), and red for full resistance (5 mutants). The *mdr1* haplotypes (N86Y/Y184F/D1246Y) are colored purple and orange to indicate resistance to AL and ASAQ, respectively. Following the shift in predominant clusters after 2014, resistance against SP decreased from partial-full (green-red) to light-partial (blue-green). Additionally, the proportion of the AL-resistant haplotype (purple) in *mdr1* haplotypes emerged after 2014.

Two predominant clusters, C61 and C62, accounted for 17 and 20% of the total samples, respectively, and persisted for 4-5 years, with C61 dominating before 2014 and C62 dominating after 2014 (Fig. 3a and 4c). Upon closer examination, C61 comprises two subgroups and C62 consists of four subgroups. The samples linking these subgroups were all polyclonal, with MOI values ranging from 2.8 to 4.1 and 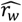 values ranging from 0.26 to 0.5 (Fig. S5a). The intra-subgroup 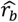 was very high (mean 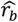 = 0.99), contrasting with the very low inter-subgroup 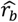 (mean 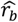 = 0.03; Fig. S5b). This indicates that the subgroups were largely unrelated, suggesting minimal effective recombination among them. The connector samples among subgroups were likely superinfections, which occurred infrequently and did not produce sufficient recombinants that circulated within the population to be frequently observed. When we lowered the 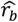 cutoff to 0.6, clear substructures remained within C61 and C62, and C61 and C62 became interconnected through few samples that were polyclonal (Fig. 4c). This, along with the low average relatedness between most clusters (Fig. 4d) and the bimodal distribution of within-year 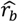 (Fig. 4e), once again suggests that the parasite clusters in STP were loosely related, with limited yet present opportunities for superinfections and effective recombination, due to low transmission intensity. Moreover, consistent with evidence of limited effective recombination, the temporal changes in drug resistance haplotypes were mainly driven by the frequency changes of different subclusters to which they belong (Fig. S5c).

In addition to temporal changes in the parasite population, we also observed spatial substructure (Fig. 4f). Parasites in or near the capital (AG and LO), where most cases occurred, exhibited similar levels of relatedness within and between districts (average 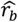 = 0.06-0.09). Conversely, two districts with lower incidence rates than the capital, Me-Zochi and Cantagalo, displayed the highest relatedness among themselves (average 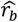 = 0.13-0.15), but lower relatedness with other districts in Sao Tome Island (average 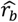 ≤ 0.09). While some general spatial substructure was evident, many clusters comprised samples from different districts, including Me-Zochi and Cantagalo, suggesting parasite flow across the entire region (Fig. S6). Interestingly, the two predominant clusters, C61 and C62, also exhibited different spatial distributions, with the former mainly existing in AG and MZ, while the latter primarily in AG and LO (Fig. 4f). With the predominant clusters changing from C61 to C62 in 2014, the number of cases in MZ also greatly decreased (Fig. S1a), suggesting limited genetic diversity in MZ.

### The role of importation and the decreased parasite diversity in the end of the study

Fig. 3 illustrates annual turnover in the parasite population, with old clusters disappearing and new clusters emerging. From 2011 to 2015, an average of 45% of clusters (12% of infections) were newly detected each year, representing a substantial amount. The newly emerged clusters may result from newly imported parasites, recombination among pre-existing parasites, and/or an increase in the frequency of infections from previously unsampled clusters. If effective recombination had occurred, we would expect to observe a group of recombinants connecting clusters. However, even after decreasing the 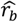 cutoff to 0.3, the subgroups were connected by few connector samples consistent with superinfections (Fig. S7). This suggests a limited role for recombination and a higher likelihood of importation. Additionally, PCA analysis revealed that samples from STP mainly clustered together with samples from Central and West Africa, situated between the clearly separated patterns of East and West African samples (Fig. 6), suggesting parasite flow between STP islands and other countries in continental Africa.

**Fig. 6.**
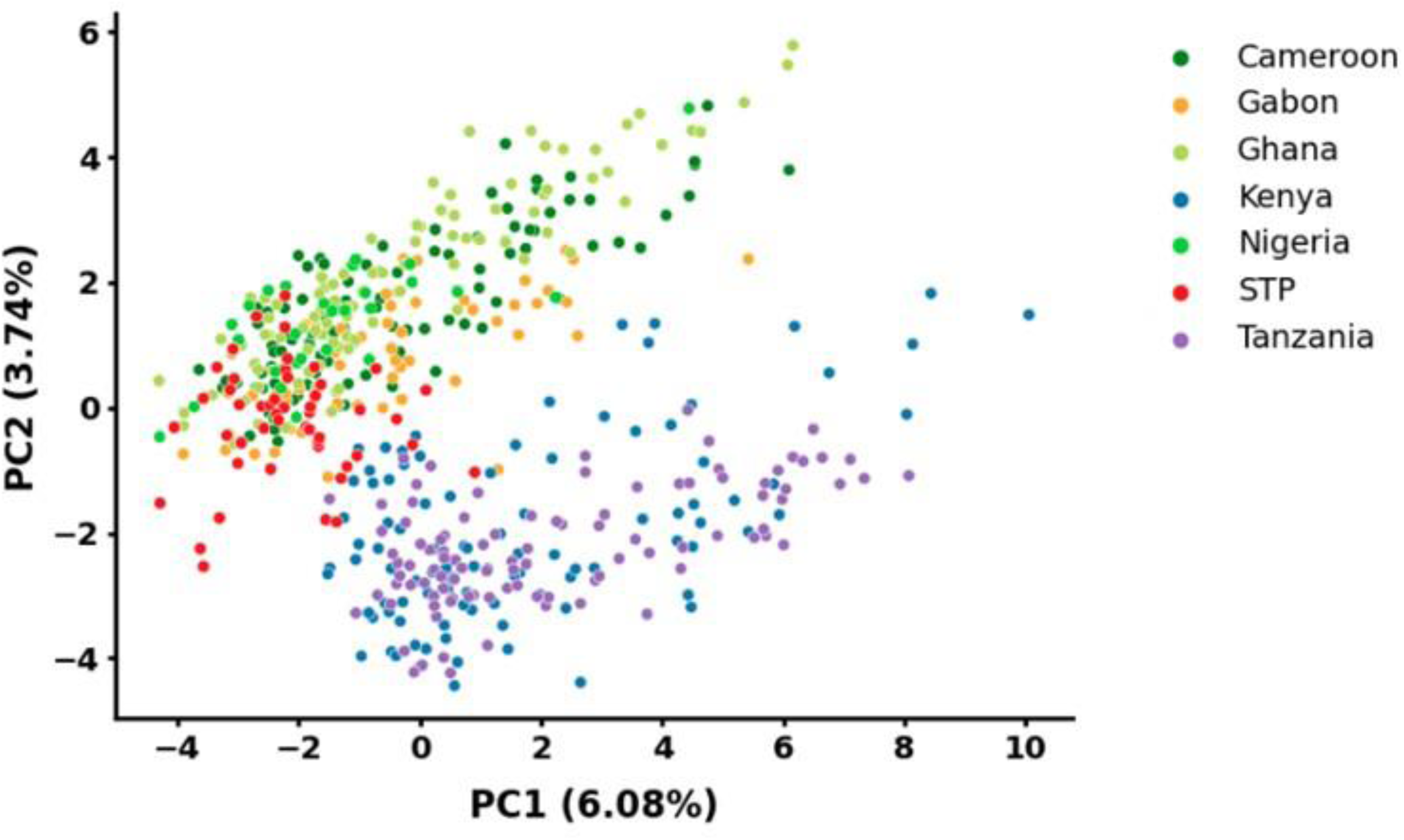
Principal component analysis (PCA) of samples from STP and six other African countries. Samples from STP are closely clustered with those from Central and West Africa.

We observed a decline in parasite diversity in the end of the study, reflected by both the number of clusters and parasite effective population size (Fig. 3a and S4). The number of clusters decreased during this period, suggesting that many clusters detected in previous years were either eliminated or reduced in size to undetectable levels (Fig. 3a). The proportion of clusters that were no longer detected in the following years increased after 2014 (Fig. 3b). While newly identified clusters were substantial from 2011 to 2015, a shift was seen in 2016, with no new clusters identified (Fig. 3c). The proportion of non-clustered or uniquely-clustered samples in 2016 was the lowest (7%) across the years, with 74% of the samples belonging to the predominant cluster C62. These data suggest a decrease in the rate of importation and/or the rates at which imported cases were successfully propagated locally in 2016.

## Discussion

Our study presents the first genomic description of the malaria parasite population in STP using amplicon sequencing technology. We offer a comprehensive depiction of the parasite population dynamics across time and space, capturing the majority of parasite genetic diversity in the country. Our findings reveal sustained, low level local transmission over a span of 7 years with moderate turnover of the parasite population likely due to sporadic propagation of imported cases. Given the consistently limited population diversity observed in STP, extending malaria genomic surveillance could serve as a valuable resource for identifying potential cases imported into STP in the future.

Genetic analysis offers valuable insights into parasite population dynamics and the effectiveness of intervention policies, complementing traditional surveillance data such as incidence of malaria ^15, 17, 22, 48, 49, 50, 51^. Incidence data may be influenced by health-seeking behaviors, and asymptomatic infections often go undetected. Furthermore, while incidence data provide changes in case numbers over time, they do not reveal the underlying structure of transmission, which is relevant when planning interventions. Our study shows that the majority of infections in STP are closely related to others, suggesting a limited number of parasite lineages circulating in STP. Moreover, while the incidence rate remained similar from 2014 to 2016, genetic analysis revealed that parasite genetic diversity was the lowest in 2016. This decline may be due to the enhanced malaria control interventions in 2015 and 2016, such as improvements in case management ^6^ and the use of new vector control methods, including outdoor larval control and the use of alternative insecticide in Indoor Residual Spraying (IRS) ^1^.

After years of malaria control, transmission intensity in several countries or regions, including STP, has decreased to a low level, with the next goal being malaria elimination ^52, 53^. In low transmission settings, genetic surveillance plays a crucial role in identifying imported cases ^54^, yet questions remain regarding the accuracy of genetic metrics in reflecting changes in underlying transmission levels ^20^. In our study, genetic metrics support the notion of low transmission intensity, but their changes over time, including the proportion of polyclonal infections, MOI, effective MOI, and effective population size, were inconsistent with changes in the incidence rate. While MOI is influenced by transmission intensity, which dictates rates of superinfection and co-transmission, it is also affected by other factors, including host age and immunity, population diversity, importation, and the genetic diversity of the locations from which imported cases originate ^55, 56^. Effective population size estimated from allele frequency changes is also influenced by importations, which increase fluctuations in allele frequencies and lead to underestimates in effective population size. In low transmission settings, genetic metrics are highly influenced by stochastic fluctuations in the parasite population. Therefore, caution is warranted when interpreting genetic metrics, and they are better considered in the context of other surveillance data and known or suspected drivers of transmission. Due to the low transmission intensity in STP, the parasite population is particularly susceptible to chance effects, and the continuous emergence of new clusters suggests the impact of importation on genetic metrics. Similarly, recent studies in Senegal and Zambia have also found a limited association between genetic metrics and incidence rates in areas with low transmission intensity ^20, 22^.

To establish the genetic basis of malaria parasites in STP, efforts were made to evenly sample across time and space, aiming to reduce bias and capture as many different genetic lineages as possible (Fig. S1). We also redefined the boundaries between years based on the transmission season to better reflect changes in malaria transmission from season to season (Fig. S3). Utilizing 165 diversity amplicons, we successfully captured both temporal changes and spatial substructure, including the continuous turnover of parasite clusters as well as genetic differentiation between some districts. Additionally, we observed a higher proportion of polyclonal infections outside the capital compared to inside, despite the higher case numbers in the capital. These polyclonal infections from non-capital districts suggest higher rates of transmission leading to superinfection, which provide opportunities for recombination and potentially increase selection efficiency ^57^. Given the connectivity between the capital and other districts (Fig. S6), these infections have the potential to spread into the capital, thereby complicating malaria control efforts.

Furthermore, evidence of a moderate degree of importation was inferred from the clustering patterns and PCA results. Despite the Sao Tome and Principe islands being approximately 300 kilometers apart from the mainland of Africa, the increased international tourism and frequent trading with high malaria transmission countries like Cameroon, Nigeria, and Gabon in recent years ^7^ make it reasonable to expect importation. However, the absence of comprehensive travel surveys has made it difficult to identify imported or introduced cases ^58, 59^. Although this study utilized genetic relatedness to understand importation, it could not fully distinguish previously unsampled parasites present in the population from imported parasites. Therefore, incorporation of a systematic travel survey, routine reactive case detection, and more comprehensive genomic surveillance into the elimination program could improve monitoring and help design and monitor strategies to reduce the impact of importation ^60, 61, 62, 63^.

Our study also characterized changes in drug resistance haplotypes of three genes, *mdr1*, *dhfr*, and *dhps* in STP, revealing a decrease of SP-resistant haplotypes and an increase in AL-resistant haplotypes. Sulfadoxine-pyrimethamine (SP) is used solely for intermittent preventive treatment (IPT) in pregnancy in STP [2] and its infrequent use may explain the decrease in SP resistance. According to WHO guidance ^64^, AL, one of the artemisinin-based combination therapies (ACT), is used as a second-line drug in STP ^6^. Among patients in our data, another ACT, AQ, is more frequently used and AL is rarely used. Thus, the increase in resistance to AL is unlikely driven by its use in STP, but is more likely led by the co-occurrence of AL-resistant and SP-lightly-resistant haplotypes in the same subgroups (Fig. S5c) and/or other non-selective factors, such as frequency changes of subgroups due to genetic drift or the emergence of new subgroups due to importation. While AL is rarely used in STP, it is recommended for treating uncomplicated infections in nearby countries such as Angola, Gabon, Nigeria, and Cameroon ^5^. Treatment failure of AL for uncomplicated *P. falciparum* malaria has been increasingly reported in sub-Saharan African countries ^65, 66, 67, 68^. Since the frequency of drug-resistant parasites can change rapidly once they evolve in a low-transmission area, routine surveillance of drug-resistance mutations or haplotypes is crucial in STP.

The last mile for STP to eliminate malaria could be challenging, given the increasing number of cases recorded in the past three years ^5, 69^. Our study revealed that the majority of malaria population was infected by highly clonal parasites with sustained transmission, while a minority were attributed to genetically distinct parasites likely originating from external sources. Therefore, we recommend disrupting the sustained local transmission by strengthening targeted interventions in common foci, and additionally implementing routine reactive surveillance and collecting travel histories, particularly for highly-mobile individuals. Lastly, this study also demonstrates how genomic surveillance can enhance our understanding of transmission dynamics. This is particularly important for low-transmission countries like STP, as it enables the establishment of strong early warning systems and aids in achieving the elimination goal.

## Conclusion

Leveraging both genomic and epidemiological data allows us to capture fine-scale dynamics of the parasite population in a pre-elimination setting. In a low-transmission setting, parasites could have experienced significant changes in population structure and drug resistance, as our study demonstrated. Targeted interventions should not only be strengthened in common foci to eliminate prevalent strains but also involve reactive case detection and tracking of travel history to prevent imported transmission in STP.

## Data Availability

All data produced in the present work are contained in the manuscript.

https://github.com/hhc-lab/malaria_genetics_STP

## Author contributions

HHC, YAC, BG, and KHT conceptualized the study. RC, LFT, and KHT led the data and sample collection. YAC and CSL designed and supervised the experiments. YAC, PYN, and AE performed experiments. YAC, DG, and PYN conducted the analysis. YAC and BP assisted with the bioinformatic analysis. YAC, HHC, and BG contributed to the interpretation of the results. YAC and HHC wrote the manuscript with help from PYN, DG, CSL, and BG.

## Acknowledgements

We would like to express our gratitude to the study participants and study teams for their cooperation, especially the Taiwan Anti-Malarial Advisory Mission and the Centro Nacional de Endemias (CNE) in STP. We also want to express our deepest gratitude to the leader of the Taiwan Anti-Malarial Advisory Mission team, Dr. Jih-Ching Lien, and to Dr. Arlindo Vicente de Assunção Carvalho from the local health ministry. Dr. Jih-Ching Lien (1927-2022) was a leading entomologist who made significant contributions to the identification of mosquito specimens. Dr. Arlindo Vicente de Assunção Carvalho (1961-2024) not only provided professional suggestions for the study but also assisted in coordinating with local departments. We thank Maxwell Murphy and Inna Gerlovina from the UCSF EPPIcenter for their comments on data analysis, Andres Aranda-Diaz for his guidance on the amplicon sequencing experiment and bioinformatic pipeline, and Ju-Hsuan Wang and Yu-Wen Huang for their help with data collection. This research was funded by the Taiwan Ministry of Foreign Affairs and the National Science and Technology Council (NSTC 113-2636-B-007-006). YAC was supported by the Fulbright Program and the UCSF EPPIcenter. HHC was supported by Yushan Scholar Program. BG was supported by NIH/NIAID mentoring award K24 AI144048.

