## Supplementary figures and tables for "Genetic surveillance reveals low, sustained malaria transmission with clonal replacement in Sao Tome and Principe"

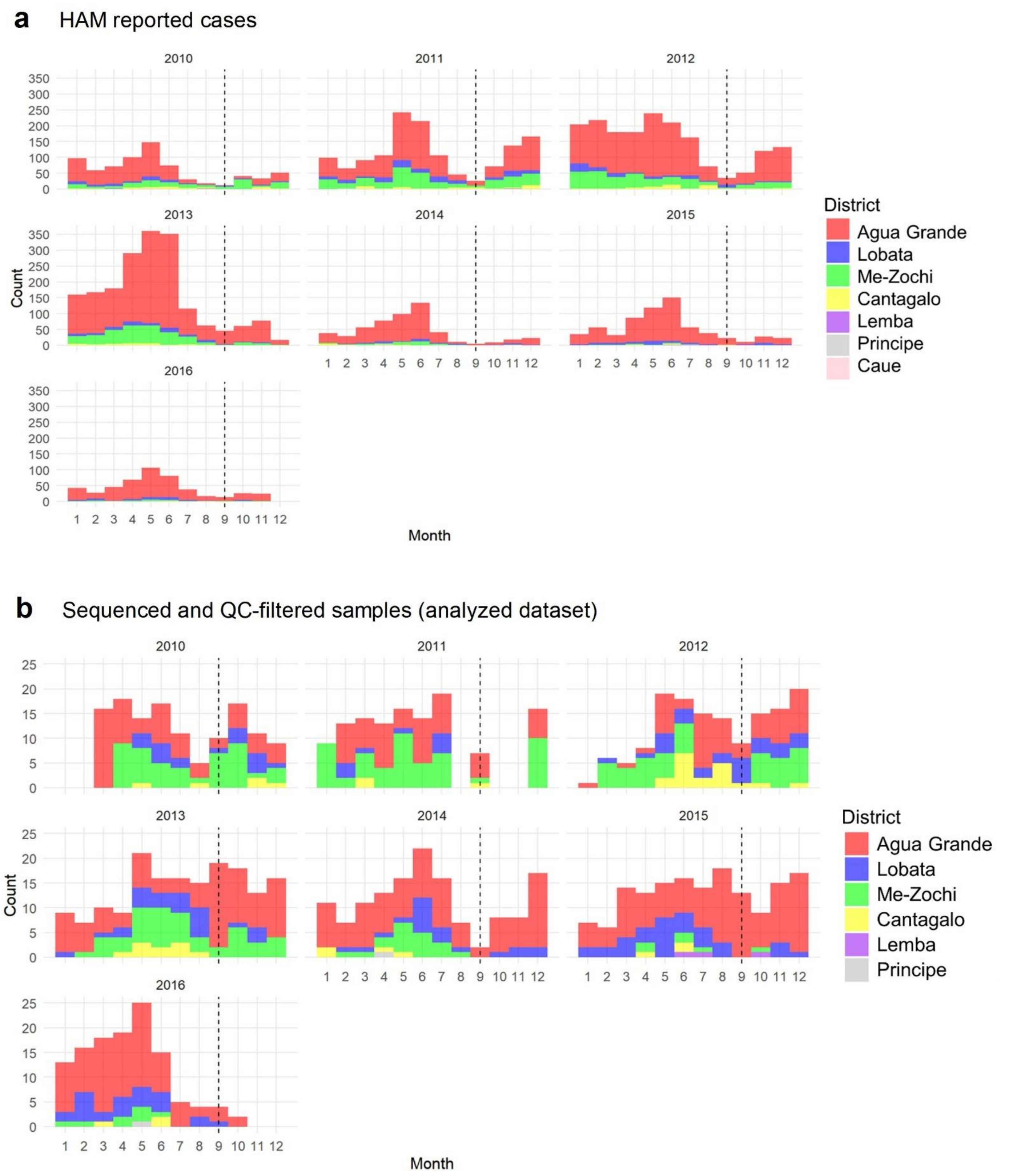


**Fig. S1. Spatial-temporal distribution of (a) total reported cases from the Central Hospital HAM, and (b) 980 sequenced samples that passed the quality check (QC) in this study.** The intervals between the dashed lines represent the reclassified years in our study. Malaria samples from the Caue district (CU), as well as samples from August, October, and November 2011, were absent in our sequenced dataset.

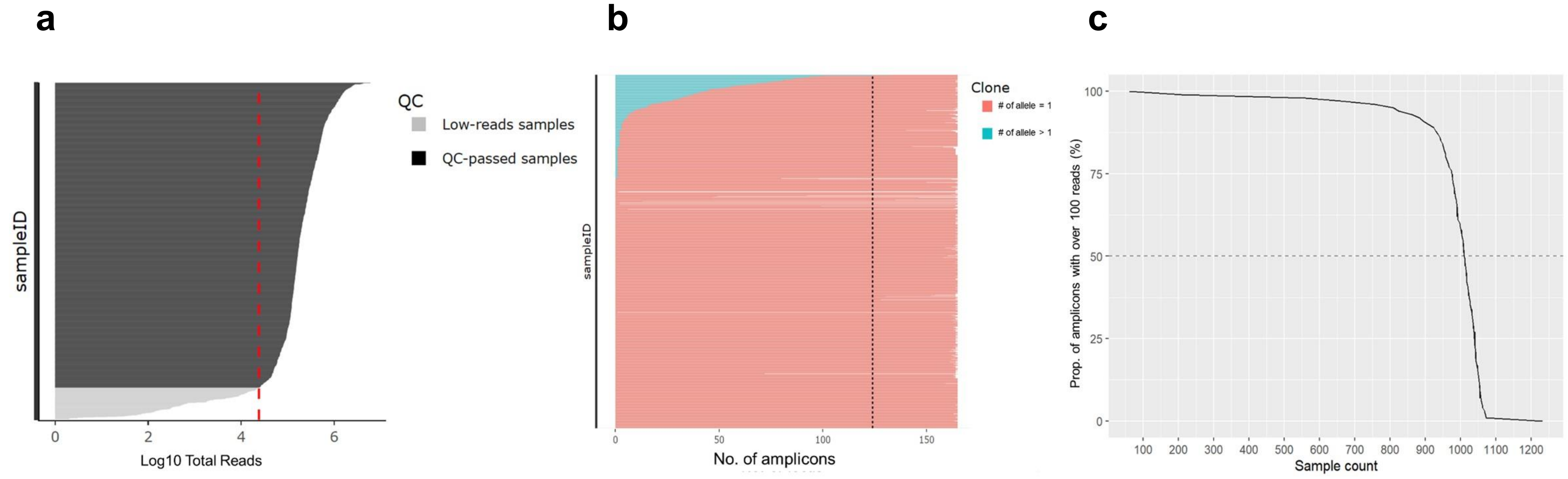


**Fig. S2. Quality check of amplicons in sequenced samples.** A total of 1,478 qPCR-positive samples were sequenced. Three criteria were employed to retain high-quality samples (*n* = 980) for diversity-related analysis, and the dashed lines in the three plots represent the filtering thresholds: (a) Total reads exceeding 100 times the number of total amplicons (about $10^{4.4}$). (b) Diversity amplicon coverage over 75% (more than 123 amplicons have non-zero reads). Colors denote the number of amplicons with more than one allele (blue), with a single allele (red), or with no information (white). (c) Over half of the diversity amplicons with the number of reads ≥ 100 (dashed line).

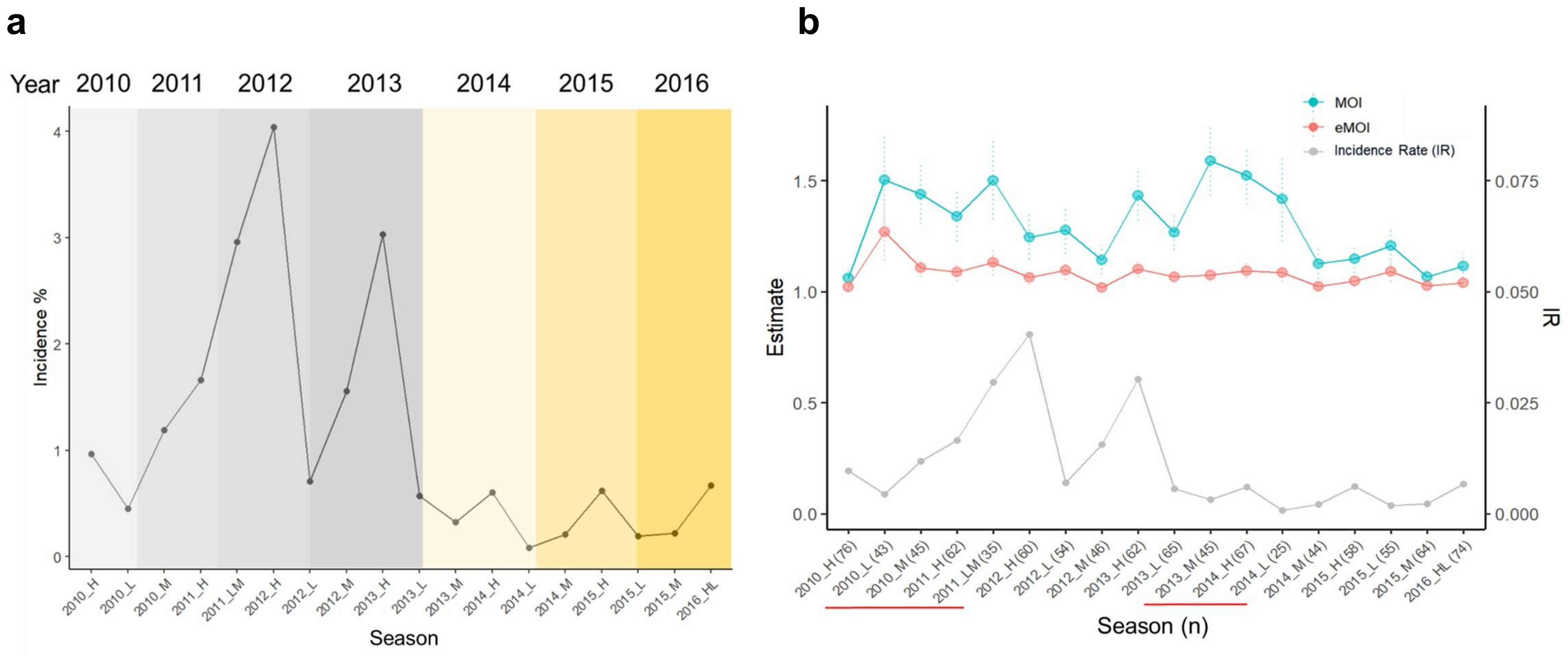


**Fig. S3. Seasonal changes of incidence rates (%), MOI, and eMOI.** (a) Transmission seasons were classified as "High (H)," "Low (L)," and "Medium (M)," corresponding to the periods from April to July, August to November, and December to March in the following year, respectively. In 2011 and 2016, two transmission seasons were combined due to the small size of sequenced samples (≤ 10). The low-transmission season in 2011 was combined with the following medium-transmission season and named 2011_LM, and the low-transmission season in 2016 was combined with the prior high-transmission season and named 2016_HL. In STP, the malaria incidence rate starts to increase around December, peak around the end of the rainy season (May), and decrease during the dry season (June to September). The redefined year (shown at the top) in this study mainly exhibited a complete peak, with the exception of 2011, during which the overall incidence rate continuously increased, as well as the beginning (2010) and the end (2016) of the study period. (b) The changes in MOI and eMOI across seasons were minimal. However, slight increases in MOI were observed in 2010-2011 and 2013-2014 (highlighted by red lines).

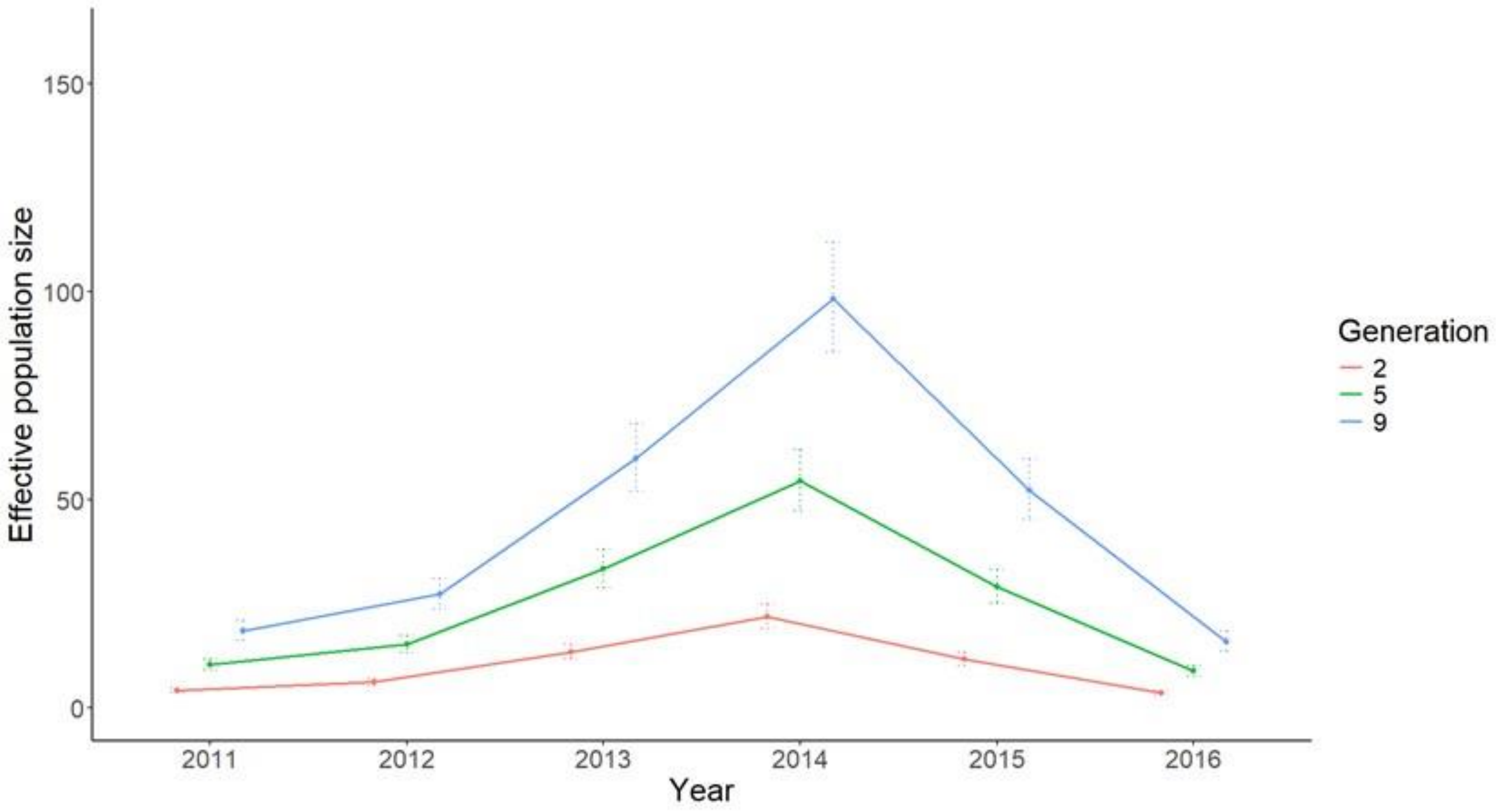


**Fig. S4. The estimated effective population sizes through time.** A range of values from 2 to 9 generations per year were explored. The error bars represent the 95% confidence intervals.

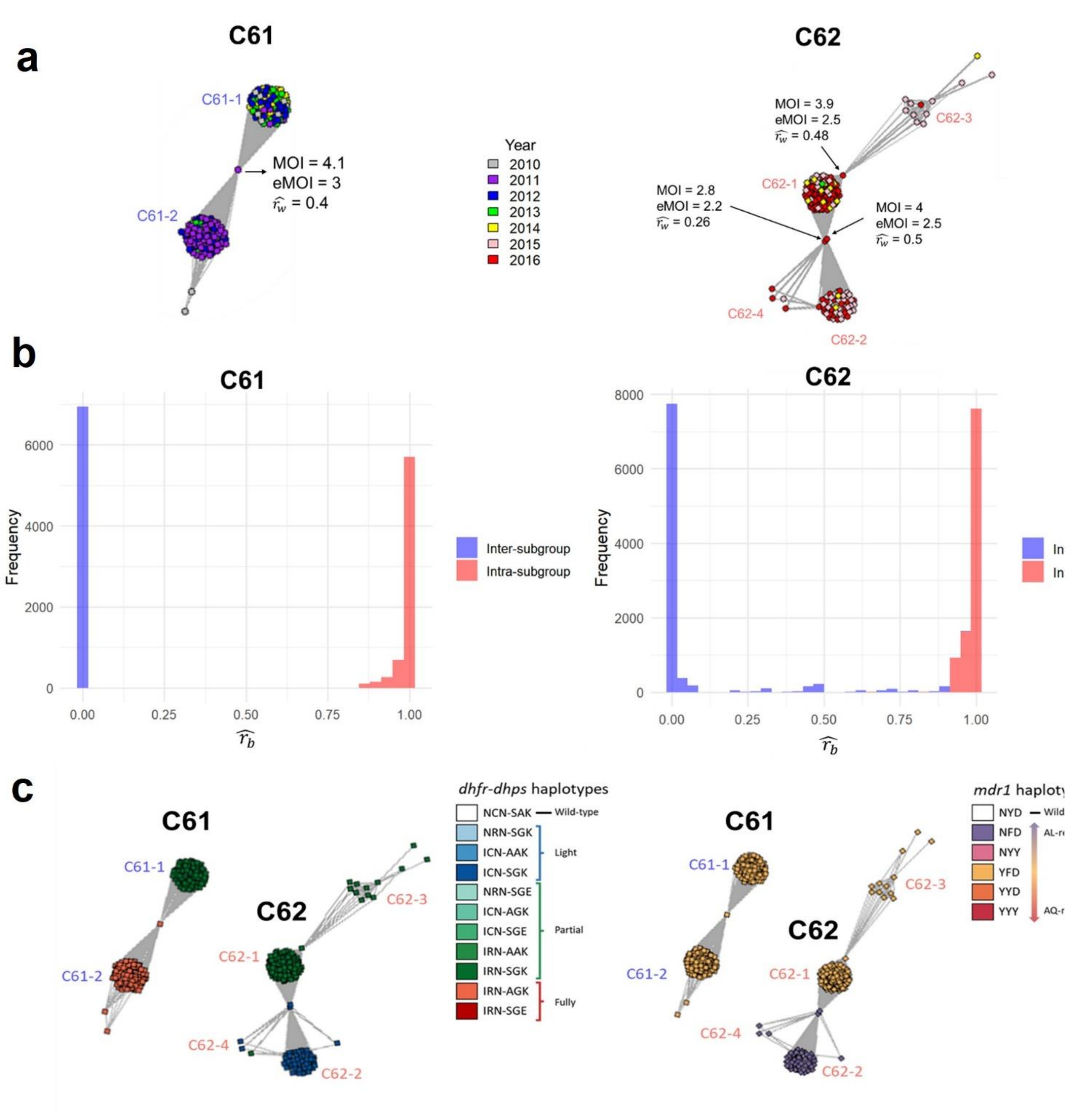


**Fig. S5. Subgroups within clusters C61 and C62.** (a) The connector nodes (samples) were labeled with MOI, eMOI, and $\widehat{r_w}$. (b) The distribution of $\widehat{r_b}$ within and between subgroups. The $\widehat{r_b}$ between connector samples and other samples was excluded from the distribution. (c) The association between drug resistance haplotypes and subgroups. The left and right graphs show *dhfr-dhps* haplotypes and *mdr1* haplotypes within subgroups of C61 and C62, respectively.

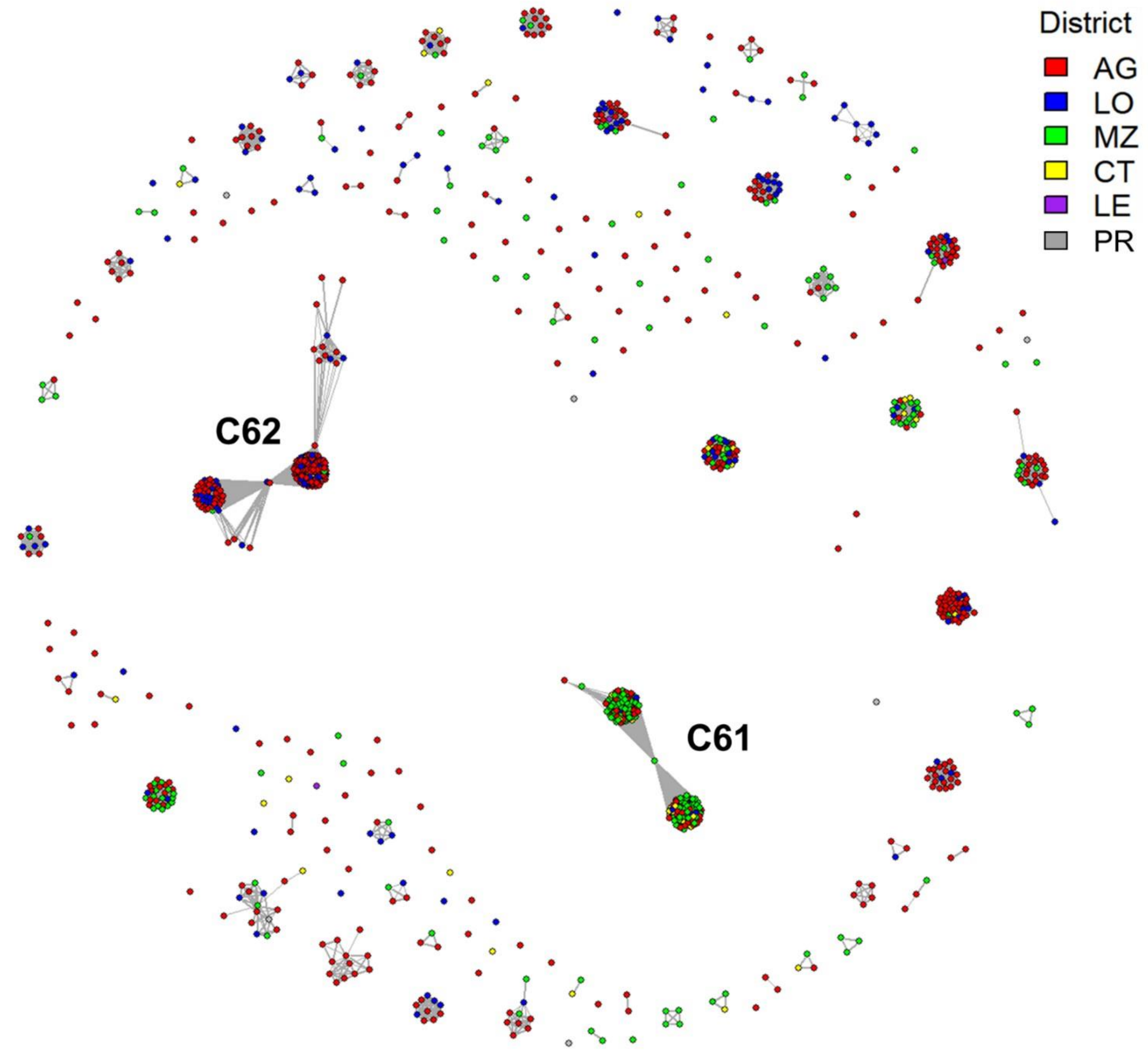


**Fig. S6. The geographical composition of districts for the clusters identified by using the $\widehat{r_b} \geq$ 0.9 cutoff.** Samples from different districts frequently clustered together.

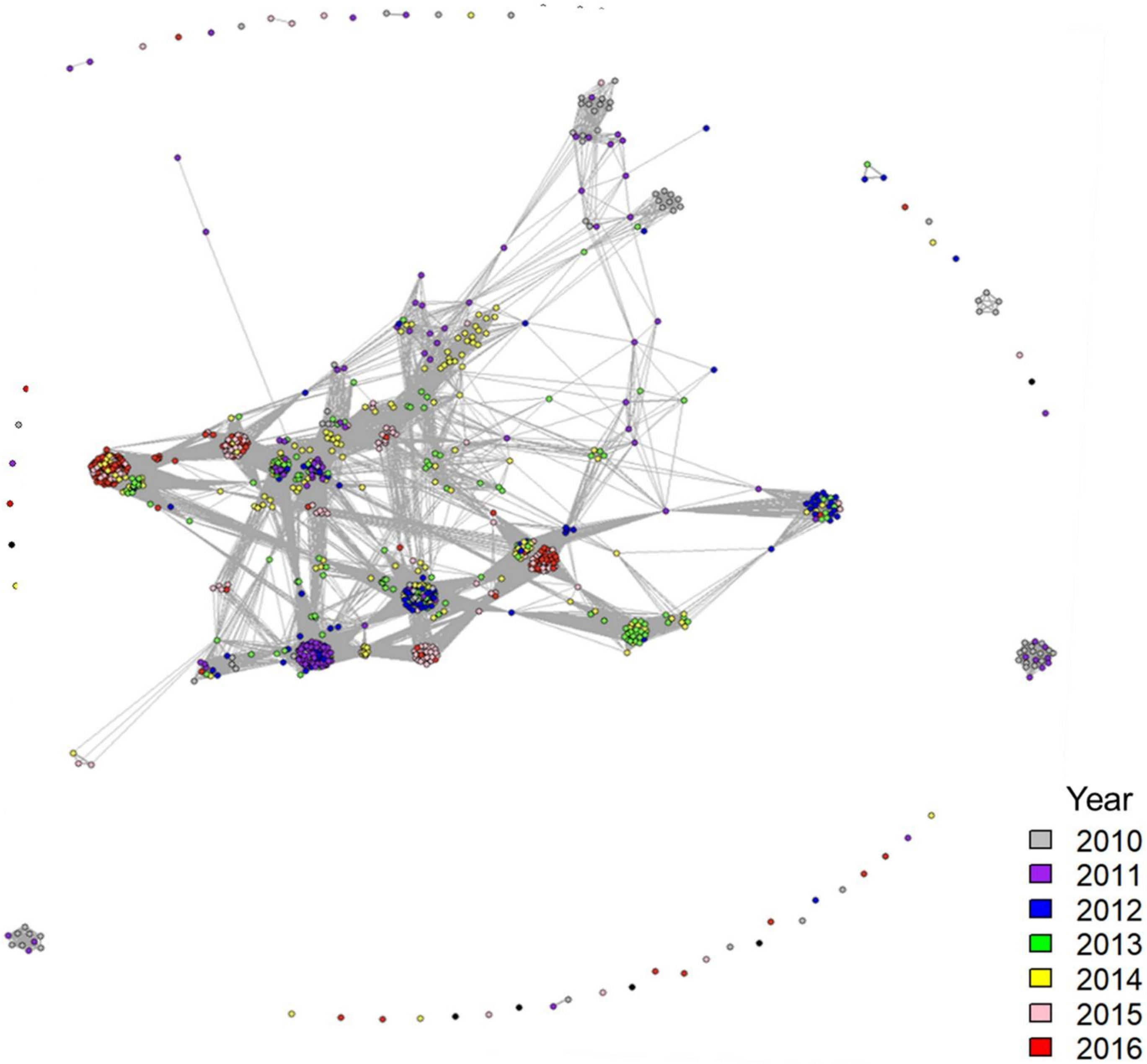


**Fig. S7. The network structure using a low cutoff of $\widehat{r_b} \geq 0.3$.** The subgroups tended to be connected by few connector samples.

**Table S1. New classification of year and area groups along with sample sizes in each group**

| Year specified in this study | Duration | n | Area |
|---|---|---|---|
| 2010 | 2010/03~2010/08 | 81 | Capital = 44<br>Others = 37 |
| 2011 | 2010/09~2011/07 | 145 | Capital = 59<br>Others = 86 |
| 2012 | 2011/09~2012/08 | 109 | Capital = 42<br>Others = 67 |
| 2013 | 2012/09~2013/08 | 163 | Capital = 64<br>Others = 99 |
| 2014 | 2013/09~2014/08 | 169 | Capital = 111<br>Others = 58 |
| 2015 | 2014/09~2015/08 | 138 | Capital = 93<br>Others = 45 |
| 2016 | 2015/09~2016/10 | 175 | Capital = 132<br>Others = 43 |

Durations were expressed in the format yyyy/mm, with underlined entries indicating different start and end points due to the lack of samples. Samples were further classified into two area groups based on their reported residential districts: "Capital" for those residing in the capital district (Agua Grande), and "Others" for those residing outside the capital district.

**Table S2. Drug-resistance mutations [1]**

| Gene | Chromosome | Amino acid position | Wild type | Mutant type |
|---|---|---|---|---|
| *dhfr* | 4 | 51 | N | I |
| *dhfr* | 4 | 59 | C | R |
| *dhfr* | 4 | 108 | S | N |
| *dhps* | 8 | 436 | S | A |
| *dhps* | 8 | 437 | A | G |
| *dhps* | 8 | 540 | K | E |
| *mdr1* | 5 | 86 | N | Y |
| *mdr1* | 5 | 184 | Y | F |
| *mdr1* | 5 | 1246 | D | Y |

**Table S3. Factors associated with MOI, eMOI, and polyclonal infection**

| Variables | MOI | | | | eMOI | | | | Polyclonal infection[#] | | | | |
|---|---|---|---|---|---|---|---|---|---|---|---|---|---|
| | Coefficient | SE | t value | p-value | Coefficient | SE | t value | p-value | Coefficient | SE | t value | p-value | OR (95%CI) |
| **Non-capital area** | 0.11 | 0.05 | 2.04 | **0.04*** | 0.06 | 0.02 | 2.74 | **0.006**** | 0.06 | 0.02 | 2.76 | **0.006**** | **1.06 (1.02, 1.11)** |
| **Location (Q1)[†]** | 0.13 | 0.06 | 2.09 | **0.04*** | 0.05 | 0.02 | 2.20 | **0.03*** | 0.07 | 0.02 | 2.90 | **0.004**** | **1.07 (1.02, 1.12)** |
| **Age above 6** | 0.17 | 0.07 | 2.54 | **0.01*** | 0.04 | 0.02 | 1.73 | 0.08 | 0.07 | 0.03 | 2.58 | **0.01**** | **1.07 (1.02, 1.13)** |
| Log10 parasite density | -0.02 | 0.03 | -0.64 | 0.53 | -0.01 | 0.01 | -0.52 | 0.60 | 0.003 | 0.01 | 0.22 | 0.82 | 1.00 (0.98, 1.03) |
| Gender (Male) | 0.06 | 0.05 | 1.27 | 0.20 | 0.01 | 0.02 | 0.31 | 0.76 | 0.03 | 0.02 | 1.31 | 0.19 | 1.03 (0.99, 1.07) |
| ACT | -0.08 | 0.06 | -1.46 | 0.15 | -0.02 | 0.02 | -1.07 | 0.29 | -0.02 | 0.02 | -0.89 | 0.37 | 0.98 (0.94, 1.03) |
| Year (2010 as reference) | | | | | | | | | | | | | |
| **2011** | 0.31 | 0.11 | 2.83 | **0.005**** | 0.11 | 0.04 | 2.79 | **0.005**** | 0.08 | 0.04 | 1.98 | **0.05*** | **1.09 (1.00, 1.18)** |
| **2012** | 0.30 | 0.11 | 2.63 | **0.009*** | 0.09 | 0.04 | 1.98 | **0.05*** | 0.11 | 0.05 | 2.40 | **0.02*** | **1.12 (1.02, 1.22)** |
| 2013 | 0.17 | 0.11 | 1.64 | 0.10 | 0.03 | 0.04 | 0.81 | 0.42 | 0.07 | 0.04 | 1.75 | 0.08 | 1.08 (0.99, 1.17) |
| **2014** | 0.40 | 0.11 | 3.85 | **0.0001***** | 0.06 | 0.04 | 1.63 | 0.10 | 0.13 | 0.04 | 3.21 | **0.001**** | **1.14 (1.05, 1.24)** |
| 2015 | 0.06 | 0.11 | 0.50 | 0.62 | 0.02 | 0.04 | 0.44 | 0.66 | 0.02 | 0.04 | 0.52 | 0.60 | 1.02 (0.94, 1.12) |
| 2016 | 0.06 | 0.11 | 0.52 | 0.61 | 0.04 | 0.04 | 0.90 | 0.37 | 0.02 | 0.04 | 0.59 | 0.55 | 1.02 (0.94, 1.11) |
| Rainy season | 0.06 | 0.05 | 1.07 | 0.28 | 0.03 | 0.02 | 1.37 | 0.17 | 0.007 | 0.02 | 0.34 | 0.73 | 1.01 (0.97, 1.05) |

[#] Polyclonal infection was assessed using logistic regression, while MOI and eMOI were assessed through linear regression.

[†] Location (Q1) denotes the hot spots with malaria case numbers ranking in the top 25% within each district. Significant factors (p ≤ 0.05) are in bold type.
